# Treatment and prevention of early disease before and after exposure to COVID-19 using hydroxychloroquine: A protocol for exploratory re-analysis of age and time-nuanced effects: Update based on initial dataset review

**DOI:** 10.1101/2020.08.19.20178376

**Authors:** David M. Wiseman, Pierre Kory, Dan Mazzucco, Mayur S. Ramesh, Marcus Zervos

## Abstract

**BACKGROUND:** Three companion randomized pragmatic trials were recently published assessing the effect of hydroxychloroquine (HCQ) on pre- exposure prophylaxis (PrEP - “Rajasingham,” NCT04328467), post- exposure prophylaxis (PEP - “Boulware,” NCT04308668) and treatment of early COVID-19 (“Skipper,” NCT04308668). Respectively, they found non-statistically significant reductions in development or persistence of COVID of 17%, 27% and 20% and concluded that HCQ did not reduce, prevent or substantially treat COVID-19 illness.

With a likely Type 2 error and over ambitious powering (50% effect), these effects may impact trajectory and resource models that drive decisions on lockdowns. All three studies have signals mostly found in the data appendices that suggest useful effects of HCQ in sub-groups worthy of re-analysis and prospective exploration. HCQ appeared to benefit younger rather than older patients in both the PrEP and PEP studies with respective reductions in COVID-19 of 45% and 36%. There was a strong benefit (40%) of HCQ in women (PrEP). Response to HCQ appeared to vary by type of exposure, with a large benefit (64%) in first responders (PrEP) and in household contacts (31%, PEP). Further confounding the data was the undefined, ex-protocol use of zinc and ascorbic acid.

**INTERIM FINDINGS:** A major impediment in interpreting the PEP and treatment studies concerns the estimation of the time from exposure or symptom onset to treatment. Our initial analysis of the PEP study as published revealed a negative correlation between treatment lag and disease reduction, reaching 49% when HCQ was initiated within one day (RR 0.51, CI 0.176-1.46, p=0.249). However, our initial review (pursuant to this protocol, v1.1) of the publicly released PEP dataset revealed that, contrary to the study’s conclusion, this four-day period referred not to the time from exposure to treatment as we (and others) had understood, but to the time from exposure and enrollment, a difference of up to 3.5 days. Our re-stratification of new data we had requested revealed that HCQ may reduce the development of COVID-19 by as much as 65% (RR 0.35, CI 0.13-0.93, p=0.044) when received within 3 days of exposure (RR 0.83 at 3-5 days; RR 1.37 at 5-7 days). There remains ambiguity in these estimates addressable by further data we have requested.

This same issue appears shared by the Treatment study. Further, patients in the PEP study were likely exposed to a series, rather than a single, “index” exposure, an issue possibly shared with the PrEP study. In the treatment study, there may be a bimodal effect of responders and non-responders, in whom symptoms may actually worsen. All three studies share the confounding effect of a possibly active folate placebo.

**OBJECTIVES:** To conduct a *post hoc* exploratory re-analyses of the de-identified raw datasets from randomized studies of the use of HCQ for pre- and post-exposure prophylaxis, and treatment of early of COVID-19 with view to further defining: (a) The time dependent effect of HCQ, on post exposure prophylaxis and treatment of COVID-19; (b) The age dependent effect of HCQ, on pre- and post- exposure prophylaxis and treatment of COVID-19; (c) The sub-stratification of gender, time- and age-dependent effects by exposure type and risk level, as well as by the use of zinc and ascorbic acid; (d) The design of prospective clinical trials designed to test the hypotheses generated by this study.

These analyses will be expanded should datasets from similarly designed Spanish studies involving PEP or treatment of (both NCT04304053) COVID-19, with directionally similar results, become available.

This protocol was devised using the Standard Protocol Items: Recommendations for Interventional Trials (SPIRIT) incorporating the WHO Trial Registration Data Set.

Study Status:

Protocol version 1.2 (September 27 2020): registered at: OSF Registries September 27 2020

https://osf.io/fqtnw

https://doi.org/10.17605/OSF.IO/FQTNW

First submitted to medrxiv 9/30/20 as version1.2a, with format revisions requested by medrxiv

Supplemental file: Detailed background and rationale

Wiseman2020Synechion2001-v1-2aReanalysisHydroxychloroquine092720medrxivREV2-SUPPL.pdf

**Plain Language Summary:** Three companion clinical studies from the University of Minnesota examined the effect of hydroxychloroquine (HCQ) when given to treat early cases of COVID-19 or to prevent it from occurring either before or just after exposure to someone with coronavirus. Although reductions in COVID-19 between 17% and 27% were found when HCQ was used before or just after exposure to COVID-19, because the studies had too few patients in them and because they were designed to find larger differences, these effects were not statistically significant, even though, they may have been clinically meaningful. Furthermore, larger reductions of COVID-19 were found within sub-groups, reaching for example in one study 45% for younger patients (<40), 40% in women and 64% in first responders. Confounding the data was the unplanned use of zinc and vitamin C.

In our initial review of the publicly released data from one of the studies, we discovered an issue that fundamentally alters its interpretation as well as one of its companion studies. According to their published reports, and understood by many others citing this work (including NIH), treatment with HCQ began within four days of either exposure or onset of symptoms. Because shipping schedules were not considered, this delay could have been 7.5 days and many patients may not have received drug in time to have an effect, if there was one. After we requested additional data, we found that HCQ may reduce COVID-19 by as much as 65% when given within 3 days of exposure and we have requested more data to clarify this figure further.

We plan therefore to conduct a re-analysis of all three studies to explore how the effects of HCQ, if any, might be dependent on time, age, gender, and type of exposure to COVID-19. This will enable us to conduct clinical studies to confirm or refute what we think we have learned from this work.

## 1 STUDY INFORMATION

### 1.1 Title

Treatment and prevention of early disease before and after exposure to COVID-19 using hydroxychloroquine: A protocol for exploratory re-analysis of age and time-nuanced effects: Update based on initial dataset review.

### 1.2 Authors

*Responsible for protocol and analysis*

David M. Wiseman PhD, MRPharmS, Synechion, Inc., Dallas, TX.

*Responsible for review of protocol, analysis*

Mayur S Ramesh MD. Henry Ford Hospital, Detroit, MI

Pierre Kory, MD. Advocate Aurora Critical Care Service, Aurora St. Luke’s Medical Center, Milwaukee, WI

Dan Mazzucco, PhD. ZSX Medical, LLC, Philadelphia, PA. Adjunct Professor, Biomedical Engineering, Rowan University, NJ

Marcus Zervos MD. Henry Ford Hospital, Detroit, MI

No professional writers will be employed.

### 1.3 Description

#### 1.3.1 Rationale

This protocol involves the exploratory re-analysis of three companion studies examining the effect of HCQ in pre-^1^ (PrEP) and post-^2^ (PEP) exposure prophylaxis, as well as Treatment^3^ of early COVID-19 in North America.

This work is being performed to clarify a number of questions arising from:

- The original publication of the post-exposure prophylaxis (PEP)^2^ study that prompted our previously registered version 1.1 of this protocol.
- The publicly released dataset for the PEP study. Our initial review of this dataset has identified a key issue not obvious in the original publication which requires this protocol revision. Further, additional clarifying data has been requested from the original authors before we can proceed with our analysis.
- The two companion studies for pre-exposure (PrEP) prophylaxis^1^ and treatment^2^ of early COVID-19. Given that these are companion studies, a number of issues are shared between them^4^ (Table 1 of Supplement). Accordingly, re-analysis of these two studies has been added in this protocol revision (1.2).

**Table 1:** Sample Sizes of Studies of Interest

| Study | N HCQ Arm | N Control Arm |
| --- | --- | --- |
| Post exposure prophylaxis<sup>2</sup> | 414 | 407 |
| Pre exposure prophylaxis<sup>1</sup> | 989 (two doses) | 494 |
| Treatment of early COVID-19<sup>3</sup> | 212 | 211 |
| Post exposure prophylaxis (Spanish)<sup>12</sup> | 1116 | 1198 |
| Treatment of early COVID-19 (Spanish)<sup>13</sup> | 136 | 157 |

A fourth companion study,^5^ not the subject of this protocol, examined the safety aspects of the other three studies and concluded: *“randomized clinical trials can safely investigate whether hydroxychloroquine is efficacious for COVID-19*.*”*

A full discussion of the issues that have provided the rationale for this re-analysis is given in the Supplemental file. In summary these issues requiring further exploration are:

##### Revised understanding of the time from exposure to treatment necessitating re-analysis

Based on a comment from the original principal investigator to us in early correspondence that there may be a “Day 1” effect shown in the original supplemental data for the PEP study, we and others^6^ found a statistically significant negative correlation (Figure 1 of Supplement) between treatment lag and reduction of COVID-19, reaching 49% when given within one day after exposure (RR 0.51, CI 0.176-1.46, p=0.249). Based on the wording of the paper itself, our initial understanding that the data described the effects of HCQ given up to four days after exposure to COVID-19 was shared by a number of others^6-11^

Pursuant to the earlier registered version (1.1) of this protocol, as we familiarized ourselves with the publicly released dataset, we discovered that this four-day period did not take into account the time to ship study medication, adding up to 3.5 days to the time lag (Table 2 of Supplement). In response to our requests, the authors provided new data to address this issue, at least partially. We re-stratified the dataset and found that HCQ may reduce the development of COVID-19 by as much as 65% (RR 0.35, CI 0.13-0.93, p=0.044) when received within 3 days of exposure (Table 3 of Supplement) with a declining effect thereafter (RR 0.83 at 3-5 days; RR 1.37 at 5-7 days). Other ambiguity in estimating the treatment lag (see Supplement) should be addressable by further data we have requested. Stratification by time is an essential element in understanding this study and must be performed before other subgroup analyses can proceed.

**Table 2:** Summary of patients forming “Responding Population” for Boulware study

|  | Total | HCQ | Placebo |
| --- | --- | --- | --- |
| Original ITT Cohort | 821 | 414 | 407 |
| *Excluded will be subjects:* |  |  |  |
| Withdrew Consent | 8 | 4 | 4 |
| LTF, no survey data per Boulware Table S1 | 55 | 25 | 30 |
| LTF, noted as "Some Survey Data", but no symptom | 9 | 5 | 4 |
| **Totals for exclusion** | **72** | **34** | **38** |
| **Total included in Responding Population** | **749** | **380** | **369** |

A similar issue appears to exist for the Treatment study^3^ which may account for the apparent bimodal effect (“improvers” vs. “non-improvers”) we discerned in the published data^4^ which suggest that since only 24% and 30% of participants accounting for the 14-day VAS of 1.5 (HCQ) and 1.87 (placebo) respectively, average scores in still-symptomatic patients *increased* to 6.15 and 6.14 in the two groups respectively.

##### Time bias related to excluded subjects

There may be time-related biases related to the exclusion of 100 randomized PEP study subjects who became symptomatic before study drug was received.

##### Age-based subgroups

Because of the need for time-based re-stratification, reductions in COVID-19 in younger subjects of similar orders of magnitude in the PEP and PrEP studies (Figure 1 Supplement) require sub-stratification. Because different age categories were used in the PrEP study than in the PEP and Treatment studies, age strata should be matched.

##### Gender-based subgroups

A reduction in COVID-19 found for women in the PrEP study (HR 0.60, CI 0.36-0.99, p=0.051) requires exploration for time and age interaction.

##### Type and level of exposure to COVID-19

There were also differences in development of COVID-19 by type of exposure (HCW vs. household in the PEP study; First Responders in the PrEP study) worthy of further exploration.

##### Observational use of Zinc and Vitamin C

The PEP and Treatment studies included the ex-protocol use of zinc and Vitamin C. Re-analysis will attempt to clarify the contribution of these agents. In the Treatment study,^3^ zinc or Vitamin C alone may have been somewhat effective without HCQ.^4^

##### Use of folate for placebo

Further confounding the study is the use of folate for the placebo which may have either inhibitory of enhancing effects on COVID-19 (Supplement). The PEP study included Canadian subjects who used a lactose placebo, subjects not taking study medication, and subjects lost to follow up or withdrawing consent. Adjusting for these subjects, we generated “folate only placebo” and “no folate control” cohorts, with a possibly lower risk of COVID-19 in the “no folate control” cohort than the folate placebo cohort (RR0.93, 95%CI 0.54-1.61) (Table 5 of Supplement). Since folate was used in all three studies, this possible effect of folate will be considered in the sub-group re-analyses for all three studies.

#### 1.3.2 Overall Aims, Objectives, Methods

##### Overall Aims

Accordingly, the now publicly released and revised dataset for the PEP study as well as the (as yet unreleased) datasets for the related and overlapping PrEP and mild disease Treatment studies may reveal important clues to inform an age-, time- and gender nuanced approach to COVID-19 using hydroxychloroquine testable by prospective studies. Data from a two similarly designed post-exposure^12^ or early treatment^13^ studies conducted in Spain (both NCT04304053), with directionally similar results may soon become available for similar analyses.

Further examination of these data may also provide insight into the various proposed mechanisms of HCQ. At early stages, consistent with incubation period estimates of 3-8 days^14^, HCQ alone may be effective in pre-emption because it interferes with viral attachment and initial infectivity. Using zinc (or Vitamin C) may be futile, ineffective or counterproductive in otherwise healthy individuals with no zinc dysregulation. HCQ may be sufficient in young people, however once infection has occurred, HCQ’s actions as an ionophore^15^ may be more important, and require the presence of zinc for viral inhibition.^16^

##### Objectives

To conduct more detailed analyses of the raw datasets relating to the use of HCQ for pre- and post-exposure prophylaxis, and treatment of early of COVID-19 with view to further defining:

a. The time dependent effect of HCQ, on post exposure prophylaxis and treatment of COVID-19.
b. The age dependent effect of HCQ, on pre- and post-exposure prophylaxis and treatment of COVID-19.
c. The sub-stratification of gender, time- and age-dependent effects by exposure type and risk level, as well as by the use of zinc and ascorbic acid.
d. The design of future clinical trials designed to test the hypotheses generated by this study.

##### Methods

A *post hoc* exploratory re-analysis will be conducted of a dataset obtained from randomized, controlled studies on the use of hydroxychloroquine to treat or prevent COVID-19. This re-analysis will be expanded should datasets from two similar Spanish studies become available. The re-analysis will explore apparent exposure risk, age and time-nuanced effects of HCQ with three main hypotheses given below.

This protocol was devised using the Standard Protocol Items: Recommendations for Interventional Trials (SPIRIT) ^17,18^ incorporating the WHO Trial Registration Data Set^19^ which it references.

### 1.4 Hypotheses

a. That HCQ exerts a negative age-dependent effect in reducing COVID-19 when given prophylactically before or after exposure.
b. The HCQ exerts a negative time-dependent effect in reducing COVID-19 when given prophylactically after exposure, or for treatment after onset of symptoms.
c. That any gender, age- and time-dependent effects of HCQ will be further dependent on type and risk level of exposure and the use of zinc or ascorbic acid.

## 2 DESIGN PLAN

### 2.1 Study type

This study is a *post hoc* exploratory analysis of a now revised dataset obtained from an original study^2^ that was a randomized, controlled, blinded clinical trial with a pragmatic design with participants from the USA and Canada. Part of the data related to the use of zinc and ascorbic acid which were observational in nature. This study seeks to conduct similar analyses on as yet unreleased datasets from two companion studies.^1,3^ Data from similarly designed open label, early treatment^13^ or open-label, cluster randomized post-exposure prophylaxis^12^ studies conducted in Spain, if available, will be subjected to similar analyses.

### 2.2 Blinding

All three companion studies^1-3^ were double-blinded studies. The Spanish post-exposure prophylaxis study^12^ was an open-label, cluster-randomized trial. Its companion^13^ early treatment study was also open label.

### 2.3 Is there any additional blinding in this study?

Details of blinding, implementation of allocation, enrollment, assignment of interventions and unblinding may be found in the publicly-released documentation for the original studies. No further blinding will be employed for the purposes of this exploratory re-analysis.

### 2.4 Study design

The original studies were interventional studies that used COVID-19 related endpoints to compare the performance of two groups - HCQ treatment vs. or placebo-^2^ or no-^12^ treatment. The current protocol will further stratify data by age-, treatment-lag, gender, exposure type and risk level and zinc and ascorbic acid use.

### 2.5 Randomization

Details of randomization are found in the publicly-released documentation for the original studies. No further randomization will be employed for the purposes of this exploratory re-analysis. Zinc and ascorbic acid were used in two studies,^2,3^ (and possibly one other^1^) but this use was not part of the protocol and was uncontrolled. Accordingly, data concerning zinc and ascorbic acid are considered as retrospective and observational.

## 3 SAMPLING PLAN

### 3.1 Existing data

The three companion^1-3^ North American studies and two Spanish^12,13^ studies have been published and examined along with their supplementary summary statistics released at the time of publication.

The raw dataset for the first (Boulware)^2^ of these has been released and has been transmitted by email to us by the original authors and was accessed after registration of version 1.1 of this protocol. On 8/20/20 the raw dataset was downloaded from:

https://drive.google.com/drive/folders/1hQS21AN3pJk1Ehm85OdDeoUut3i-b8wt This consisted of two files named:

NEJM_Contents_DataDictionary_17July2020.csv (7kb)

NEJM_PEP_PublicDataSet_17July2020.csv (281kb)

After accessing these data, a quality control check found a number of discrepancies with the published study for tallies of several study variables. Clarification was sought from the study authors on these points as well as other aspects of the of data, most notably questions relating to:

- Calculation of the time from exposure to drug receipt (see <u>1.3</u>)
- Data from 100 randomized but excluded patients who became symptomatic before receipt of study drug.
- Identification of patients who did not take any drug, or who did not complete the course of drug.
- Use of folate as placebo
- Exposure risk based on use of PPE

Acknowledging the errors, the authors transmitted to us a revised dataset on 9/9/20 consisting of two files:

PEP_DataDictionary_09Sep2020.csv (10kb)

PEP_Public_Data_09Sep2020.csv (288kb)

Our quality control check on this revised dataset confirmed the agreement of variable tallies with those found in the published paper as well as the correction of the discrepancies previously identified. Clarification was provided regarding the use of folate and the identification of subjects fully, partially or non-adherent to the study medication. Regarding our remaining question regarding assessment of exposure risk, we were informed that the definition of exposure risk stated in the paper was incorrect and required correction. The study authors were not prepared to provide data for the excluded 100 patients as those data would be later released as part of the dataset for the companion (sharing the same NCT registration) study,^3^ although according to that publication, deidentified participant data was to be made available from July 22 2020.

Regarding the calculation of exposure to drug receipt time, a new file was transmitted containing further information about the time from enrollment to the receipt of study drug:

PEP_Exposure_to_MedArrival_time.csv (9kb)

The parameter provided was the sum of two numbers:

- The integer number of days from exposure to screening(=enrollment) (as previously provided) plus
- The actual number of hours (divided by 24) from time-stamped enrollment to time-stamped FEDEX delivery.

The calculation of this parameter was confirmed as the following example:

> A subject enrolled at 3pm on Monday and noted their days to exposure =1. The subject received drug the next morning at 10.30am (19.5 hours later) to produce a total exposure to arrival time (as provided) of 1 + 19.5/24 = 1.81 days. The exposure may have occurred just before midnight Sun-Mon in which case total time would be (15+19.5)/24 = 1.44 days, or just after midnight (Sat-Sunday) in which case the total time would be (15+24+20.5)/24 = 2.44 days. Further, since enrollment could have taken place at any time during a 24-hour period, these estimates are subject to an additional variation of ± 24 hours.

Accordingly, we renewed our request for information regarding the time (of day) of enrollment that would narrow this window for each subject and provide more accurate time stratification. Also, in response to our various questions, we were informed that for the Canadian participants, same-day courier service was used, although the delivery lag was not known. Since the particular data we requested may be protected by privacy regulations, we proposed means to de-identify it whilst preserving its value for this re-analysis.

To help determine appropriate analyses, an interim estimate of the time-stratified effect of hydroxychloroquine on development of COVID-19 was performed using these new (but incomplete) data and is given in Table 3 of the Supplement. Further, data have been examined to determine the “Responding Population” of patients who provided symptom data (<u>5.5, Table 2</u>).

Accordingly, data have been partially accessed, but additional requested data will permit more precise estimations of the time from exposure to delivery, as well as the conduct of the other analyses.

The datasets for the companion^1,3^ and Spanish studies^12^ have not been released. Accordingly, this protocol revision is being registered prior to accessing those data: As of the date of submission of this revised protocol, the data exist, but have not been accessed by the sponsor or principal investigator.

### 3.2 Explanation of existing data

We have requested additional data that will permit more precise time stratification of the data. The kinds of analyses to be performed will depend on whether this occurs and in what form.

### 3.3 Data collection procedures

Details of data collection procedures including inclusion and exclusion criteria are found in the publicly-released documentation for the original studies. No further data collection will be employed for the purposes of this exploratory re-analysis.

### 3.4 Sample size

Details of sample size are found in the publicly-released documentation for the original studies (<u>Table 1</u>).

### 3.5 Sample size rationale

Sample size rationales are provided in the original studies. This exploratory study will examine smaller subgroups and without control over their size. The statistical challenges of performing sub-group analyses are well-known.^20,21^

### 3.6 Stopping rule

Not applicable

## 4 VARIABLES

### 4.1 Manipulated variables

The manipulated variable (treatment arm) employed in the original studies is:

Treatment, or pre- or post-exposure prophylaxis of COVID-19 with levels as:

- Treatment with HCQ (two dose intervals for pre-exposure prophylaxis study)
- No (or placebo) treatment

Data will be stratified and sub-stratified by:

- Subject age and gender
- Time between exposure to COVID-19, or symptom onset and treatment.
- Type of exposure (household vs. health worker contact etc.)
- Level of exposure risk
- Treatment with zinc or ascorbic acid
- Full, partial and non-adherence to taking study drug with view to exploring the relationship between the use of folate and COVID-19. Data from Canadian subjects taking lactose placebo, will be pooled with data from subjects taking no study medication.

Strata containing small numbers of subjects will be pooled with adjacent strata as appropriate and to meet sample size requirements of the tests to be employed.

### 4.2 Measured variables

The primary outcomes defined in the original studies were:

- Presence of COVID-19, based on symptom-based criteria with expert review.^2^
- Presence of PCR-confirmed, symptomatic Covid-19^12^

Secondary endpoints were:

- Prevention of transmission of COVID-19^12^

### 4.3 Indices

The presence of symptom- or test-based COVID-19 will be expressed a percentage of the number of subjects for each subgroup and Odds or Risk Ratios computed with 95% confidence intervals. Risk ratios may also be expressed as its complement, the reduction of risk.

## 5 ANALYSIS PLAN

### 5.1 Statistical models

Fisher’s Exact or Chi Square tests will be used as appropriate to compare the incidence of COVID-19 in HCQ- and control groups, for each sub-group stratification described above. If appropriate for the nature of the requested data, univariate regression analyses will be conducted to examine the effect of age- and time lag on any effect of HCQ. The possibility will be explored of conducting multivariate Cox regression analyses with propensity score matching to examine observational data relating to the use of zinc and ascorbic acid. See also comments received after version 1.0 of this protocol was registered from Dr. Boulware (section <u>5.6</u>).

Should the dataset become available for the two Spanish^12,13^ studies, it may be possible to aggregate data with those from the respective North American studies. As this is an exploratory analysis, no adjustment will be made for the clustering used in the Spanish study.

### 5.2 Transformations

Depending on whether and what form remaining data are received concerning exposure to drug receipt time, other possible analyses and the need for data transformation cannot yet be determined.

### 5.3 Inference criteria

Since all analyses performed in this protocol are exploratory in nature, p-values from two-tailed tests will be reported if ≤ 0.1 without adjustment for multiple comparisons.^21^

### 5.4 Data exclusion

The ability to conduct the planned analyses will depend on the format of the data remaining to be supplied.

### 5.5 Missing data

An Intent-to-Treat (ITT) analysis will be employed as was done in the original studies. As this protocol is exploratory in nature, sensitivity analyses will not be conducted. Data will also be analyzed utilizing those subjects for whom endpoint (symptom) data is present, as the “Responding Population” (RP) (<u>Table 2</u>).

For the PEP study, details of patients withdrawing consent (8) or lost to follow up (88, LTF) are provided in Figure 1 and Table S1 there. Of the 88 LTF patients, 52 were reported as not completing any surveys and were unresponsive to follow up. Another 36 had: some survey data with vital status after day 14 known (16), no survey with vital status after day 14 known (3) or no survey with vital status after day 14 unknown (17). We examined the 33 patients noted as having some survey data and found that there were 9 with no symptom data. There is thus a total of 72 patients with no symptom data at all which we will exclude from the Responding Population. The remaining 24 patients had incomplete symptom data for days 3, 5, 10 and 14 in various combinations and will be included using the “last observation carried forward” (LOCF) method with the endpoint adjudication determined by the original authors.

It is anticipated that we will handle missing data from the companion studies similarly.

### 5.6 Exploratory analysis

A *per protocol* analysis will be attempted to explore the effect, if any, of the folate placebo, by segregating data from subjects according to their use of HCQ, folate, lactose or no study drug. We will construct “Folate placebo” and “No folate Control” cohorts by aggregating patients noted as taking no study medication (regardless of randomization), along with Canadian patients taking lactose placebo (Table 5 of Supplment)

Other relationships between the primary outcomes and demographic variables, as well as exposure type and level will be explored.

Subsequent to the registration (https://osf.io/4akug) of version 1.0 of this protocol, we received the following unsolicited but welcome suggestions from Dr. Boulware, the principal author of the first^2^ of the studies considered in this protocol.

a. *“Along with doing the incorrect Watanabe linear regression analysis, consider doing the actual proper analysis for categorical data, which would be a logistic regression model with an interaction term for treatment group * exposure time group*.” **Response:** The ability to conduct this type of analysis will depend on the level of data granularity which is as of this protocol version (1.1) unknown to us.
b. *“Look at the actual percentages by each of the subgroup days of exposure. There are some minor variation in the event rate between 12*.*7% to 17*.*9% between days in the placebo. As these are small subgroups, this creates artifact when looking at the difference. Consider, pooling the placebo event rate across the Day 1-4 exposure groups. If you can say why the Day 2 placebo group incidence rate would be higher than Day 1 or 3 or 4, sure keep it as is. If you pool the placebo together, the nice perfectly linear line is much less linear. For example, the day 2-3 HCQ incidence is 12% vs 12*.*2%, respectively. The placebo day 2-3 incidence is 16*.*9% vs. 14*.*5%, respectively. If you want to say day 2 works great but not day 3, that’s a bit of a stretch. Yes, the difference is 4*.*9% vs 2*.*3% --but this is an over-interpretation of the subgroup analysis – when the incidence of disease at day 2 and 3 is virtually the same with HCQ*.*”* **Response:** The suggestion to pool the placebo data may be reasonable as there really should not be any difference by time and all we are seeing there is noise. However, there is still the possibility that folate has some activity [see <u>1.3]</u>, in which case pooling may not be appropriate.

## Supporting information

Detailed background and rationale

## Data Availability

The publicly released dataset from the three main studies discussed herein by Boulware and colleagues (University of Minnesota) is/will be available at: https://covidpep.umn.edu/data

https://osf.io/fqtnw

## 6 OTHER

### 6.1 Trial identifiers and Registration

This protocol has been registered with the Open Science Framework (OSF, Center for Open Science). This study has the internal identifier of SYN2001SYN.

### 6.2 Protocol Version and Revision History

Version 1.0 8/13/20 Registered: OSF osf.io/hyp8k DOI: doi.org/10.17605/OSF.IO/4AKUG https://osf.io/fgd53/

Version 1.1 8/19/20 Registered: OSF osf.io/9rpyt DOI: doi.org/10.17605/OSF.IO/9RPYT

a. Posted at medrxiv 8/26/20 as preprint^22^
b. Registration details for v1.0 added
c. Co-authors (MR, PK, DM) added (<u>1.2</u>)
d. Minor typos and grammatical errors
e. Suggestions from Dr. Boulware (<u>5.6</u>)

Version 1.2 9/27/20 Registered: OSF https://osf.io/fqtnw DOI: doi.org/10.17605/OSF.IO/FQTNW

In reviewing the released PEP data, we discovered that shipping times (up to 3.5 days) were not considered in the published analysis. Requesting additional data, our re-stratification suggest that HCQ received within 3 days may reduce illness by as much as 65%. Before proceeding, this finding necessitates protocol revision, further data clarification, and the inclusion of companion studies sharing this and/or related issues.

<u>Version 1.2a 9/30/20 First submitted to medrxiv</u>

i. Per medrxiv request, detailed protocol rationale moved to supplement, with summary in new section <u>1.3.1.</u>
ii. Abstract and Summary shortened
iii. Correction of typos, OSF url and doi information
iv. Requested changes annotated in v1.2 revision list

Revision list for v1.2 (annotated for medrxiv changes in v1.2a)

a. Title change from: *“Preventing disease after exposure to COVID-19 using hydroxychloroquine: A summary of a protocol for exploratory re-analysis of age and time-nuanced effects*.*”*
b. Plain language summary added. Abstract revised accordingly.
c. Scope of protocol widened to include PrEP and Treatment studies, based on subject overlap and common issues between the three companion studies.^1-3^ The possible inclusion of a second^13^ Spanish study, for early treatment of COVID-19 also now added.
d. Registration details for v1.1
e. Co-author MZ added <u>(1.2</u>)
f. Relationship to Watanabe analysis.^6^ (<u>1.3,</u> now in Supplement).
g. Sub headings added for Background section. (<u>1.3</u>).
h. Discussion of findings related to treatment lag (summary in<u>1.3.1,</u> expanded in Supplement).
i. Discussion of missing data from censored subjects (summary in<u>1.3.1,</u> expanded in Supplement).
j. Revised HCW estimate from CDC (<u>1.3,</u> moved to Supplement).
k. Discussion of possible negative effects of folate as placebo (summary in <u>1.3.1,</u> expanded in Supplement).
l. Discussion of companion study on pre-exposure prophylaxis.^1^ (summary in<u>1.3.1,</u> expanded in Supplement).
m. Relationship to companion study on early treatment (summary in <u>1.3.1,</u> expanded in Supplement).
n. Revise aims, objectives etc. to include widened scope of companion studies (<u>1.4</u>).
o. Revised time stratification based on new data on shipping times (summary in <u>1.3.1,</u> expanded in Supplement).
p. First data access (8/20/20) (<u>3.1</u>)
q. Update on dataset per QC check and dataset clarifications received to 9/18/20 <u>(3.1</u>).
r. Analysis for effect of folate on COVID-19, sub stratification for gender (<u>4.1</u>).
s. Chi square test added (<u>5.1</u>)
t. Revised missing data, definition of “Responding Population” (<u>5.5</u>)
u. Per protocol analysis added (<u>5.5</u>)
v. Update privacy issues (<u>6.6</u>)
w. Record clarifying dataset: https://osf.io/tsmp6/

### 6.3 Sponsor and Contact Information

Contact for public and scientific queries:

Principal Investigator: Dr. David Wiseman

Synechion, Inc., 18208 Preston Road, Suite D9-405, Dallas, 75252

972 931 5596

There are no secondary sponsors

### 6.4 Study Schedule

The original studies have been completed and their data published.^1-3,12^

### 6.6 Ethics, Consent and Confidentiality

Analyses will be performed on a de-identified, publicly released dataset obtained from studies conducted under ethics committee supervision. No further Ethics Committee, IRB approval, informed consent or confidentiality provisions are required. We have requested additional data from the Boulware study regarding the time (of day) of enrollment. This request is being reviewed by the Privacy Officer of the University of Minnesota, along with our proposals for appropriate deidentification, if needed.

### 6.7 Roles of committees

Not applicable

### 6.8 Harms, ancillary and post-trial-care

Not applicable

### 6.9 Data management and access

No data entry is required. Microsoft Excel will be used for primary data manipulation. Vassar Stats (<u>vassarstats.net/</u>) will be used for confirmation of calculations, as well as other statistical software as appropriate. A study report will be complied and submitted for publication. Microsoft Excel files will be made available on request.to other investigators.

### 6.10 Summary Results

Data will be summarized as part of the study publication. It remains to be determined whether our re-analyses will be published together within one publication, or separately.

### 6.11 Appendix

Consent materials: Not applicable

Biological specimens: Not applicable

### 6.12 SPIRIT CHECKLIST of PROTOCOL ITEMS

The checklist for the Standard Protocol Items: Recommendations for Interventional Trials (SPIRIT) ^17,18^ was used to construct this protocol. As the SPIRIT checklist references (Item 2b) the WHO Trial Registration Data Set^19^, a cross reference is provided below for the 24-item WHO Data Set. Where there is no corresponding SPIRIT item, it is listed under SPIRIT Item 2b. Section numbers for the required item are indicated. Non-applicable items are marked “NA.”

SPIRIT 2013 Checklist: Recommended items to address in a clinical trial protocol and related documents*

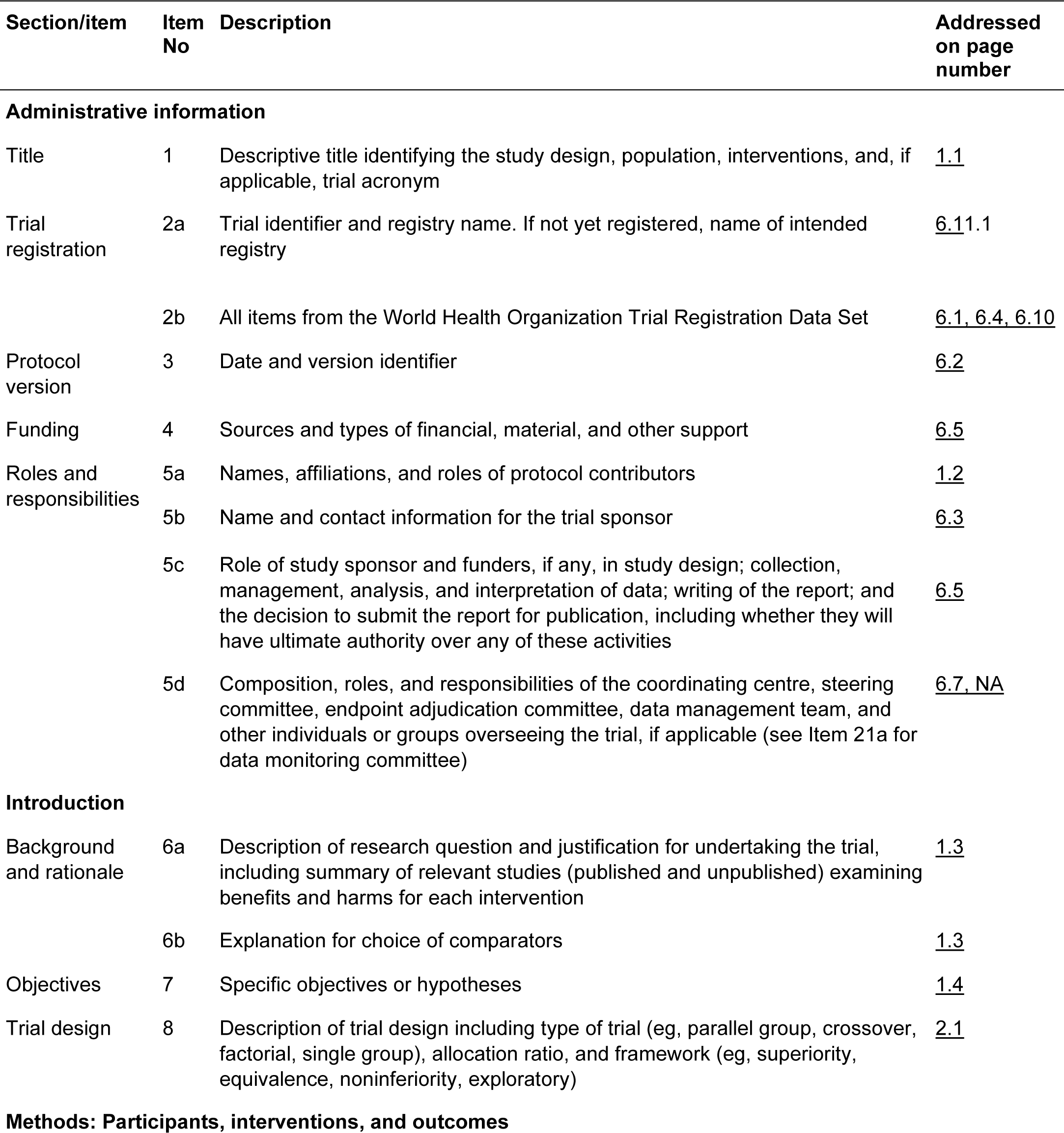

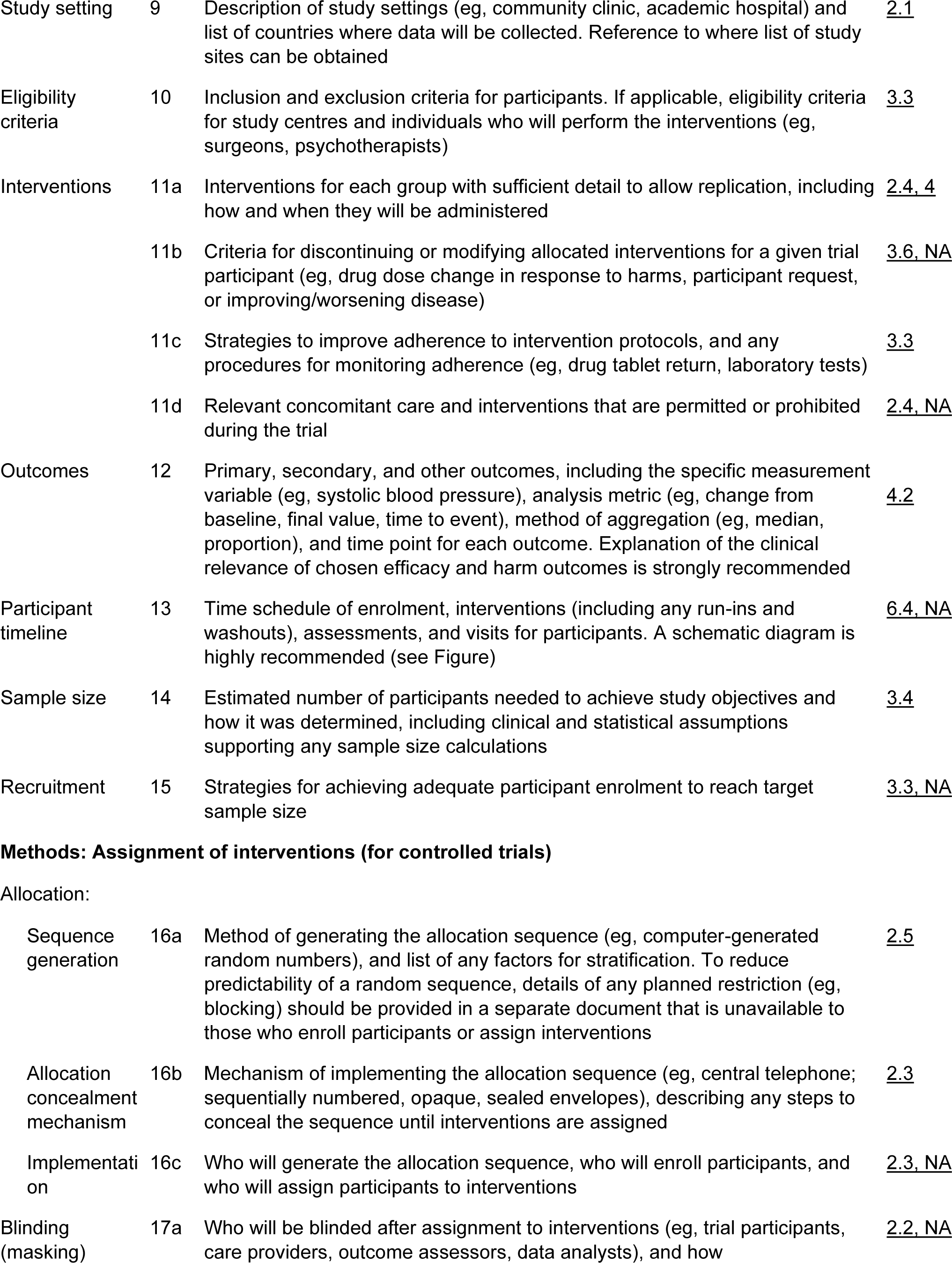

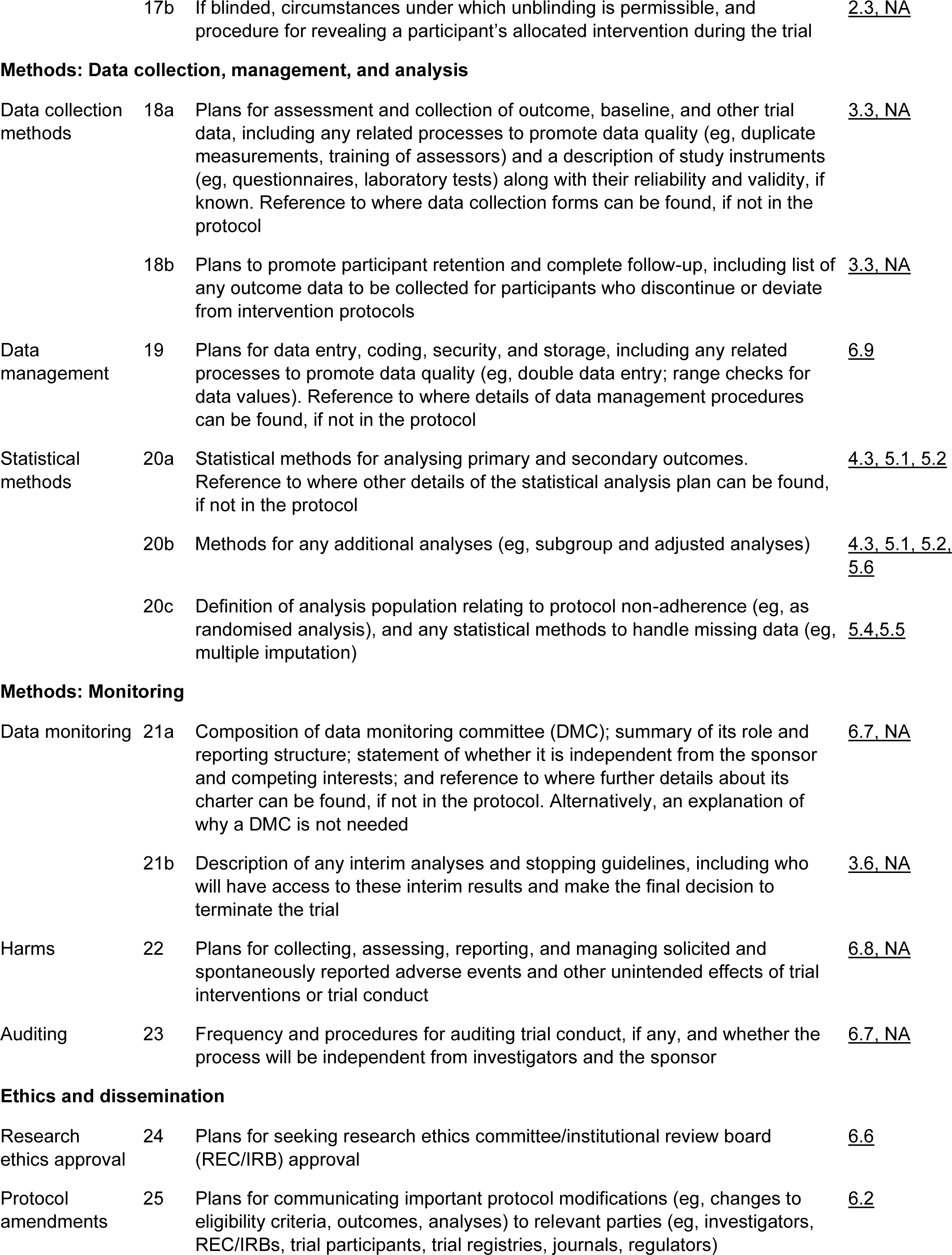

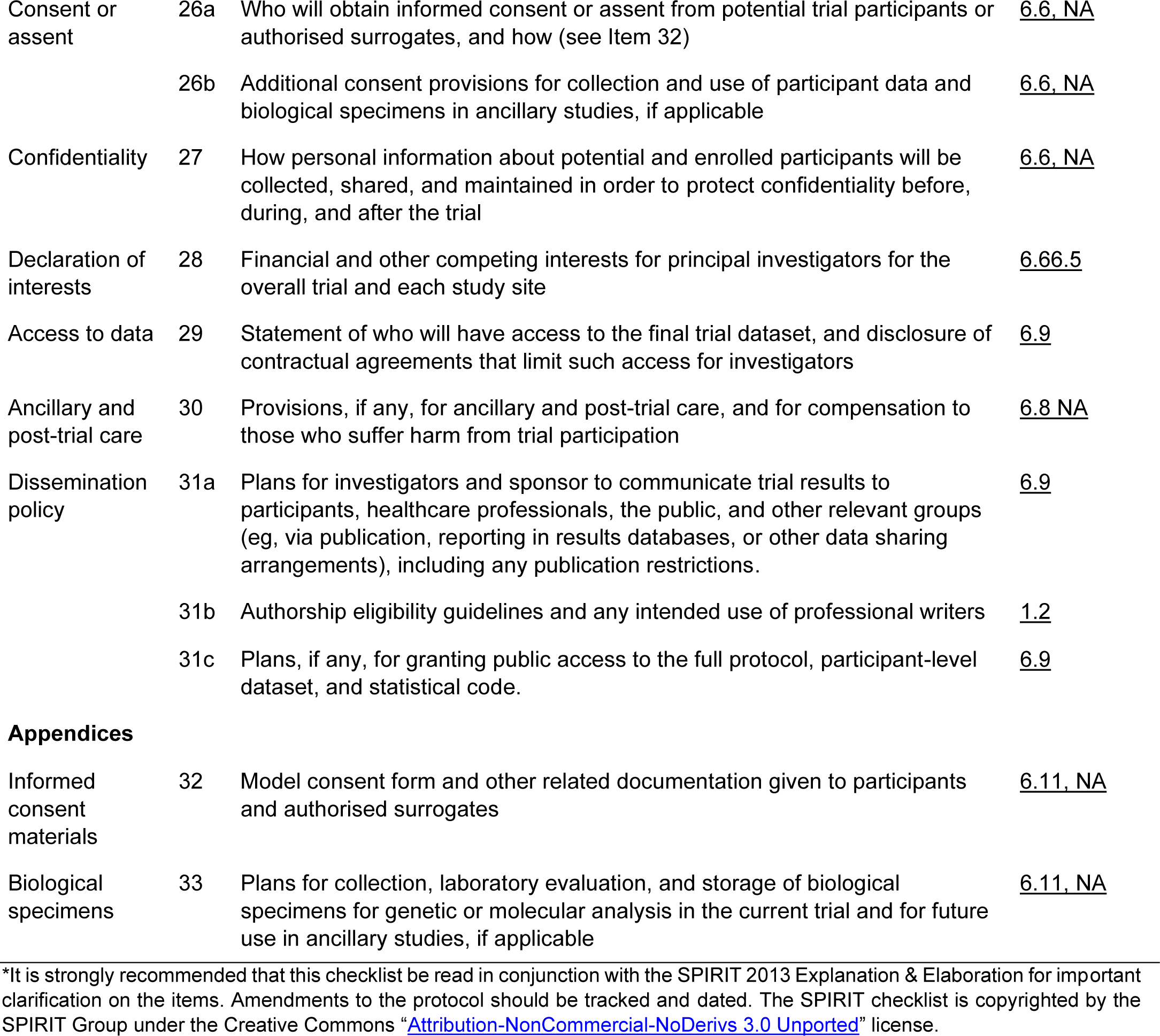

| Section/item | Item No | Description | Addressed on page number |
| --- | --- | --- | --- |
| **Administrative information** |  |  |  |
| Title | 1 | Descriptive title identifying the study design, population, interventions, and, if applicable, trial acronym | <u>1.1</u> |
| Trial registration | 2a | Trial identifier and registry name. If not yet registered, name of intended registry | <u>6.11.1</u> |
|  | 2b | All items from the World Health Organization Trial Registration Data Set | <u>6.1, 6.4, 6.10</u> |
| Protocol version | 3 | Date and version identifier | <u>6.2</u> |
| Funding | 4 | Sources and types of financial, material, and other support | <u>6.5</u> |
| Roles and responsibilities | 5a | Names, affiliations, and roles of protocol contributors | <u>1.2</u> |
|  | 5b | Name and contact information for the trial sponsor | <u>6.3</u> |
|  | 5c | Role of study sponsor and funders, if any, in study design; collection, management, analysis, and interpretation of data; writing of the report; and the decision to submit the report for publication, including whether they will have ultimate authority over any of these activities | <u>6.5</u> |
|  | 5d | Composition, roles, and responsibilities of the coordinating centre, steering committee, endpoint adjudication committee, data management team, and other individuals or groups overseeing the trial, if applicable (see Item 21a for data monitoring committee) | <u>6.7, NA</u> |
| **Introduction** |  |  |  |
| Background and rationale | 6a | Description of research question and justification for undertaking the trial, including summary of relevant studies (published and unpublished) examining benefits and harms for each intervention | <u>1.3</u> |
|  | 6b | Explanation for choice of comparators | <u>1.3</u> |
| Objectives | 7 | Specific objectives or hypotheses | <u>1.4</u> |
| Trial design | 8 | Description of trial design including type of trial (eg, parallel group, crossover, factorial, single group), allocation ratio, and framework (eg, superiority, equivalence, noninferiority, exploratory) | <u>2.1</u> |
| **Methods: Participants, interventions, and outcomes** |  |  |  |
| Study setting | 9 | Description of study settings (eg, community clinic, academic hospital) and list of countries where data will be collected. Reference to where list of study sites can be obtained | 2.1 |
| Eligibility criteria | 10 | Inclusion and exclusion criteria for participants. If applicable, eligibility criteria for study centres and individuals who will perform the interventions (eg, surgeons, psychotherapists) | 3.3 |
| Interventions | 11a | Interventions for each group with sufficient detail to allow replication, including how and when they will be administered | 2.4, 4 |
|  | 11b | Criteria for discontinuing or modifying allocated interventions for a given trial participant (eg, drug dose change in response to harms, participant request, or improving/worsening disease) | 3.6, NA |
|  | 11c | Strategies to improve adherence to intervention protocols, and any procedures for monitoring adherence (eg, drug tablet return, laboratory tests) | 3.3 |
|  | 11d | Relevant concomitant care and interventions that are permitted or prohibited during the trial | 2.4, NA |
| Outcomes | 12 | Primary, secondary, and other outcomes, including the specific measurement variable (eg, systolic blood pressure), analysis metric (eg, change from baseline, final value, time to event), method of aggregation (eg, median, proportion), and time point for each outcome. Explanation of the clinical relevance of chosen efficacy and harm outcomes is strongly recommended | 4.2 |
| Participant timeline | 13 | Time schedule of enrolment, interventions (including any run-ins and washouts), assessments, and visits for participants. A schematic diagram is highly recommended (see Figure) | 6.4, NA |
| Sample size | 14 | Estimated number of participants needed to achieve study objectives and how it was determined, including clinical and statistical assumptions supporting any sample size calculations | 3.4 |
| Recruitment | 15 | Strategies for achieving adequate participant enrolment to reach target sample size | 3.3, NA |
| Allocation: |  |  |  |
| Sequence generation | 16a | Method of generating the allocation sequence (eg, computer-generated random numbers), and list of any factors for stratification. To reduce predictability of a random sequence, details of any planned restriction (eg, blocking) should be provided in a separate document that is unavailable to those who enroll participants or assign interventions | 2.5 |
| Allocation concealment mechanism | 16b | Mechanism of implementing the allocation sequence (eg, central telephone; sequentially numbered, opaque, sealed envelopes), describing any steps to conceal the sequence until interventions are assigned | 2.3 |
| Implementation | 16c | Who will generate the allocation sequence, who will enroll participants, and who will assign participants to interventions | 2.3, NA |
| Blinding (masking) | 17a | Who will be blinded after assignment to interventions (eg, trial participants, care providers, outcome assessors, data analysts), and how | 2.2, NA |
|  | 17b | If blinded, circumstances under which unblinding is permissible, and procedure for revealing a participant's allocated intervention during the trial | <u>2.3, NA</u> |
| Data collection methods | 18a | Plans for assessment and collection of outcome, baseline, and other trial data, including any related processes to promote data quality (eg, duplicate measurements, training of assessors) and a description of study instruments (eg, questionnaires, laboratory tests) along with their reliability and validity, if known. Reference to where data collection forms can be found, if not in the protocol | <u>3.3, NA</u> |
|  | 18b | Plans to promote participant retention and complete follow-up, including list of any outcome data to be collected for participants who discontinue or deviate from intervention protocols | <u>3.3, NA</u> |
| Data management | 19 | Plans for data entry, coding, security, and storage, including any related processes to promote data quality (eg, double data entry; range checks for data values). Reference to where details of data management procedures can be found, if not in the protocol | <u>6.9</u> |
| Statistical methods | 20a | Statistical methods for analysing primary and secondary outcomes. Reference to where other details of the statistical analysis plan can be found, if not in the protocol | <u>4.3, 5.1, 5.2</u> |
|  | 20b | Methods for any additional analyses (eg, subgroup and adjusted analyses) | <u>4.3, 5.1, 5.2, 5.6</u> |
|  | 20c | Definition of analysis population relating to protocol non-adherence (eg, as randomised analysis), and any statistical methods to handle missing data (eg, multiple imputation) | <u>5.4, 5.5</u> |
| Data monitoring | 21a | Composition of data monitoring committee (DMC); summary of its role and reporting structure; statement of whether it is independent from the sponsor and competing interests; and reference to where further details about its charter can be found, if not in the protocol. Alternatively, an explanation of why a DMC is not needed | <u>6.7, NA</u> |
|  | 21b | Description of any interim analyses and stopping guidelines, including who will have access to these interim results and make the final decision to terminate the trial | <u>3.6, NA</u> |
| Harms | 22 | Plans for collecting, assessing, reporting, and managing solicited and spontaneously reported adverse events and other unintended effects of trial interventions or trial conduct | <u>6.8, NA</u> |
| Auditing | 23 | Frequency and procedures for auditing trial conduct, if any, and whether the process will be independent from investigators and the sponsor | <u>6.7, NA</u> |
| Research ethics approval | 24 | Plans for seeking research ethics committee/institutional review board (REC/IRB) approval | <u>6.6</u> |
| Protocol amendments | 25 | Plans for communicating important protocol modifications (eg, changes to eligibility criteria, outcomes, analyses) to relevant parties (eg, investigators, REC/IRBs, trial participants, trial registries, journals, regulators) | <u>6.2</u> |
| Consent or assent | 26a | Who will obtain informed consent or assent from potential trial participants or authorised surrogates, and how (see Item 32) | <u>6.6, NA</u> |
|  | 26b | Additional consent provisions for collection and use of participant data and biological specimens in ancillary studies, if applicable | <u>6.6, NA</u> |
| Confidentiality | 27 | How personal information about potential and enrolled participants will be collected, shared, and maintained in order to protect confidentiality before, during, and after the trial | <u>6.6, NA</u> |
| Declaration of interests | 28 | Financial and other competing interests for principal investigators for the overall trial and each study site | <u>6.66.5</u> |
| Access to data | 29 | Statement of who will have access to the final trial dataset, and disclosure of contractual agreements that limit such access for investigators | <u>6.9</u> |
| Ancillary and post-trial care | 30 | Provisions, if any, for ancillary and post-trial care, and for compensation to those who suffer harm from trial participation | <u>6.8 NA</u> |
| Dissemination policy | 31a | Plans for investigators and sponsor to communicate trial results to participants, healthcare professionals, the public, and other relevant groups (eg, via publication, reporting in results databases, or other data sharing arrangements), including any publication restrictions. | <u>6.9</u> |
|  | 31b | Authorship eligibility guidelines and any intended use of professional writers | <u>1.2</u> |
|  | 31c | Plans, if any, for granting public access to the full protocol, participant-level dataset, and statistical code. | <u>6.9</u> |
| Informed consent materials | 32 | Model consent form and other related documentation given to participants and authorised surrogates | <u>6.11, NA</u> |
| Biological specimens | 33 | Plans for collection, laboratory evaluation, and storage of biological specimens for genetic or molecular analysis in the current trial and for future use in ancillary studies, if applicable | <u>6.11, NA</u> |

\*It is strongly recommended that this checklist be read in conjunction with the SPIRIT 2013 Explanation & Elaboration for important clarification on the items. Amendments to the protocol should be tracked and dated. The SPIRIT checklist is copyrighted by the SPIRIT Group under the Creative Commons "Attribution-NonCommercial-NoDerivs 3.0 Unported" license.

**Cross Reference of SPIRIT Checklist and WHO Data Set**

| SPIRIT ITEM | WHO Item | WHO Description |
| --- | --- | --- |
| 1 | 10 | Scientific Title |
| 1 | 9 | Public Title |
| 1 | 12 | Health Condition(s) or Problem(s) Studied |
| 2a | 3 | Secondary Identifying Numbers |
| 2a | 1 | Primary Registry and Trial Identifying Number |
| 2b (additional items) | 2 | Date of Registration in Primary Registry |
| 2b | 18 | Recruitment Status |
| 2b | 23 | Summary Results |
| 4 | 4 | Funding |
| 4 | 5 | Secondary Sponsor(s) |
| 5b | 5 | Sponsor |
| 5b | 6 | Contact for Public Queries |
| 5b | 7 | Contact for Scientific Queries |
| 5b | 8 | Contact information |
| 8 | 15 | Description |
| 9 | 11 | Setting, counties |
| 10 | 14 | Eligibility |
| 11a | 13 | Interventions |
| 12 | 19 | Primary Outcomes |
| 12 | 20 | Secondary Outcomes |
| 13 | 16 | Date of First Enrollment |
| 13 | 22 | Completion date |
| 14 | 17 | Sample Size |
| 24 | 21 | Ethics review |
| 31c | 24 | IPD sharing statement |

