## Supplementary material for "Treatment and prevention of early disease before and after exposure to COVID-19 using hydroxychloroquine: A protocol for exploratory re-analysis of age and time-nuanced effects: Update based on initial dataset review": Detailed background and rationale

Version 1.2a 9/27/20

#### SUPPLEMENTAL FILE: Detailed background and rationale

David M. Wiseman, PhD, MRPharmS;[1] Pierre Kory, MD; [2] Dan Mazzucco, PhD;[3] Mayur S. Ramesh, MD,[4] Marcus Zervos, MD. [4]

[1] Synechion, Inc., Dallas, TX. Who

[2] Aurora St. Luke's Medical Center, Milwaukee, WI

[3] ZSX Medical, LLC, Philadelphia, PA

[4] Henry Ford Hospital, Detroit, MI

Address for correspondence:

Dr. David Wiseman, Synechion, Inc., 18208 Preston Road, Suite D9-405, Dallas, 75252


Study Status:
Protocol version 1.2 (September 27 2020)
Protocol registered at: OSF Registries September 27 2020: https://osf.io/fqtnw
https://doi.org/10.17605/OSF.IO/FQTNW

Submitted to medrxiv 9/30/20 as version1.2a, with revisions requested by medrxiv

### 1 INTRODUCTION

This protocol involves the exploratory re-analysis of three companion studies examining the effect of HCQ in pre-[1] and post-[2] exposure prophylaxis, as well as treatment[3] of early COVID-19 in North America.

The purpose of this supplementary document is to provide the rationale for our proposed re-analysis performed to clarify a number of questions arising from:

- The original publication of the post-exposure prophylaxis (PEP)[2] study that prompted our previously registered version 1.1 of this protocol.
- The publicly released dataset for the PEP study. Our initial review of this dataset has identified a number of issues not obvious in the original publication which require this protocol revision. Further, additional clarifying data has been requested from the original authors before we can proceed with our analysis.
- The two companion studies for pre-exposure (PrEP) prophylaxis[1] and treatment[2] of early COVID-19. Given that these are companion studies, a number of issues are shared between them (Table 1). Accordingly, re-analysis of these two studies has been added in this protocol revision (1.2).

A fourth companion study,[4] not the subject of this protocol, examined the safety aspects of the other three studies and concluded: *"randomized clinical trials can safely investigate whether hydroxychloroquine is efficacious for COVID-19."*

*Table 1: Apparent issues in three companion studies on the effects of HCQ in pre- and post- exposure prophylaxis and treatment of COVID-19.*

| Description/Issue | Pre-exposure prophylaxis | Post-exposure prophylaxis | Treatment of early COVID-19 |
|---|---|---|---|
| First author | Rajasingham[1] | Boulware[2] | Skipper[3] |
| Design | RCT (pragmatic?, not stated) | RCT, pragmatic | RCT, pragmatic |
| NCT Registration | NCT04328467 | NCT04308668 | NCT04308668 |
| Subjects | High exposure risk HCW | High exposure risk HCW or household contacts | Symptomatic non-hospitalized adults with PCR or symptom-based COVID-19 |
| N HCQ arm | 989 (two doses) | 414 | 212 |
| N Placebo arm | 494 | 407 | 211 |
| Dose | 800mg divided loading dose, then 400mg once or twice weekly x12 weeks | 1.4g loading dose (divided) day 1<br>600mg daily x 4 days | 1.4g loading dose (divided) day 1<br>600mg daily x 4 days |
| Placebo | Folate 1.6mg as loading dose, then 800mcg once or twice weekly x 12 weeks. (Canada 4mg as loading, 2mg once or twice weekly) | Folate 2.8mg day 1, 1.2mg daily x 4 days (lactose for Canadian patients) | Folate 2.8mg day 1, 1.2mg daily x 4 days (lactose for Canadian patients) |
| Effect size in power calculation | 50% (80% power), hampered by early termination due to poor recruitment | 50%, 90% power | 50% (90% power) reduction in hospitalization or deaths, changed to 90% power to detect difference of 0.25 points in VAS score. |
| Endpoint | COVID-19 by symptoms or PCR | COVID-19 by symptoms or PCR | • Unvalidated, left censored VAS score<br>• Symptom severity change over 14 days |
| Overall HR (95% CI) | 0.73 (0.48-1.09) p=0.12 | 0.83 (0.58-1.18) | 0.8, 0.58-1.1, p= 0.21 (for symptom persistence |
| Possible effects in subgroups HR | For combined dose groups:<br>• Age < 40 HR 0.55 (0.32 - 0.96, p=0.038)<br>• Female (HR 0.60, 0.36 - 0.99, p=0.051)<br>• First responders (HR 0.36, 0.15 - 0.88, p=0.036) | • Age <35 years HR 0.64<br>• Household contacts (RR 0.691, 0.398-1.2, p=0.24) | • Age >50 24% relative difference in VAS score<br>• Male 21%, female 4.8% relative difference in VAS score<br>• The use of the VAS score obscures an apparent population bimodality ("improvers" vs. "non-improvers"). With only 24% and 30% of subject accounting for the 14-day VAS of |

| | | | |
|---|---|---|---|
| | • Adherent to study medication (HR 0.67, CI 0.42 - 1.08)<br>• Different age categories used than for companion studies. | | 1.5 (HCQ) and 1.87 (placebo) respectively, average scores in still-symptomatic patients *increased* to 6.15 and 6.14 respectively. |
| Age categories | **Total HCQ Placebo**<br>18-39 420 232<br>40-60 505 243<br>>60 64 19 | **Total HCQ Placebo**<br>18-35 151 145<br>36-50 159 171<br>>50 104 91 | **Total HCQ Placebo**<br>18-35 69 83<br>36-50 94 78<br>>50 49 50 |
| Time from exposure | Mostly not applicable, see below | RR 0.51 < 1 day (per study report) | |
| Time stratification | • Subjects may have already been exposed to COVID-19 prior to study (sensitivity analysis indicates little impact on overall outcome)<br>• Some time bias possible due to lag in delivery of study medication, but sensitivity analysis indicates little impact on overall outcome. Overall time frame much longer than for companion studies | • Includes only time from "exposure" to enrollment. Does not include time to receipt of study drug. Critical timeframe should be extended from 4 to ~ 7 days.<br>• Subject stratification will change with more complete estimate of timeframe.<br>• With enrollment to delivery time, RR is 0.36 for drug given within <3 days of exposure. Uncertainty remains (± 24 hours) due to incomplete and non-granular data.<br>• Uncertainty regarding identification of a single "index" exposure vs. multiple exposures.<br>• Possible time bias related to 100 randomized subjects become symptomatic before receipt of study drug. | • Enrollment of patients with < 4 days of symptoms does not take into account delivery time of study medication, possibly adding up to 3 days.<br>• An unknown number of patients may have initiated therapy far too late for a possible effect.<br>• The reduction in VAS score change of 26% with a duration of symptoms of 1-2 days (score difference 0.66 (95%CI -1.29 to -0.02) considered implausible by authors may be explained by inappropriate time stratification due to failure to consider enrollment to drug delivery time. |
| Active placebo | • Folate may affect development of COVID-19, however lower and less frequent dosing than companion studies | • Folate may affect development of COVID-19 | Folate may affect development of COVID-19 |
| Zinc, ascorbic acid | • Not stated, but expected | • Undefined, ex-protocol and observational use confounding overall results | Undefined, ex-protocol and observational use confounding overall results. Possible benefit of zinc or ascorbic acid in patients not taking HCQ. |

#### 2 POST EXPOSURE PROPHYLAXIS (PEP) STUDY OF HYDROXYCHLOROQUINE

A recently published randomized trial ("Boulware") found a non-statistically significant reduction in Covid-19 of 17% when hydroxychloroquine (HCQ) was used for postexposure prophylaxis. The study concluded *"After high-risk or moderate-risk exposure to Covid-19, hydroxychloroquine did not prevent illness compatible with Covid-19 or confirmed infection when used as postexposure prophylaxis within 4 days after exposure."* [2] A number of issues prompt the exploratory re-analysis of the study described in this protocol.

*Sample Size, Powering and Clinically Meaningful Effect Size*

Based on practical considerations[5] rather than considerations of clinical meaningfulness, the study was powered to detect reduction of development of COVID-19 by 50%. Since this was a "pragmatic" clinical trial, where effect sizes are typically smaller than under tightly controlled "explanatory" trial conditions,[6] and with greater heterogeneity, this may have been over-ambitious. Based on CDC (cdc.gov/coronavirus/2019-ncov/cases-updates/cases-in-us.html, 9/11/20) underestimates of 158,519 COVID-19 cases among Health Care Workers (HCW) in the USA, a 17% reduction in COVID-19 would have translated to a reduction of about 26,948 cases. Since a similar figure of 17% was considered clinically meaningful for a reduction of mortality using dexamethasone in severe hospitalized cases of COVID-19,[7] a 17% reduction of COVID-19 in certain populations may impact modeling of COVID-19 trajectory and health resource estimates that drive decisions on lockdowns and social distancing.

*Supplemental Data Regarding Age-based Subgroups*

Supplemental data not discussed in the main body of the published paper suggested greater reductions of COVID-19 in a number of sub-groups. There were reductions (36%) in younger (<35 years) and increases (110%) in older (> 50 years) subjects (Figure 1). This is consistent with the findings of an RCT from Boulware and his colleagues on the pre-exposure prophylactic use of HCQ[1] (Figure 1), as well as an observational prophylaxis study involving mainly younger HCW in India.[8] Luco[9] has performed an independent re-analysis of these data and has also concluded that there may be an age-dependent benefit to HCQ.

*Supplemental Data Regarding Time from Exposure*

Based on a comment of Dr. Boulware in early private correspondence regarding a possible “Day 1” effect found in the supplemental data, we conducted a preliminary analysis that revealed a statistically significant negative correlation (Figure 1) (slope -0.211, 95%CI -0.328—0.094, p=0.016, rho CI -1 to -0.42, confirmed independently[10 a]) between treatment lag and reduction of COVID-19, reaching 49% when given within one day after exposure (RR 0.51, CI 0.176-1.46, p=0.249). The early use of HCQ is supported by mathematical modeling that considers the peaking of the viral load in the pharyngeal cavity and the effect of the drug to kill infected cells by enhancing cell-mediated immunity.[11]

Figure 1: *Prophylaxis with HCQ: Time Lag (Post) and Age Dependency (Pre/post)*

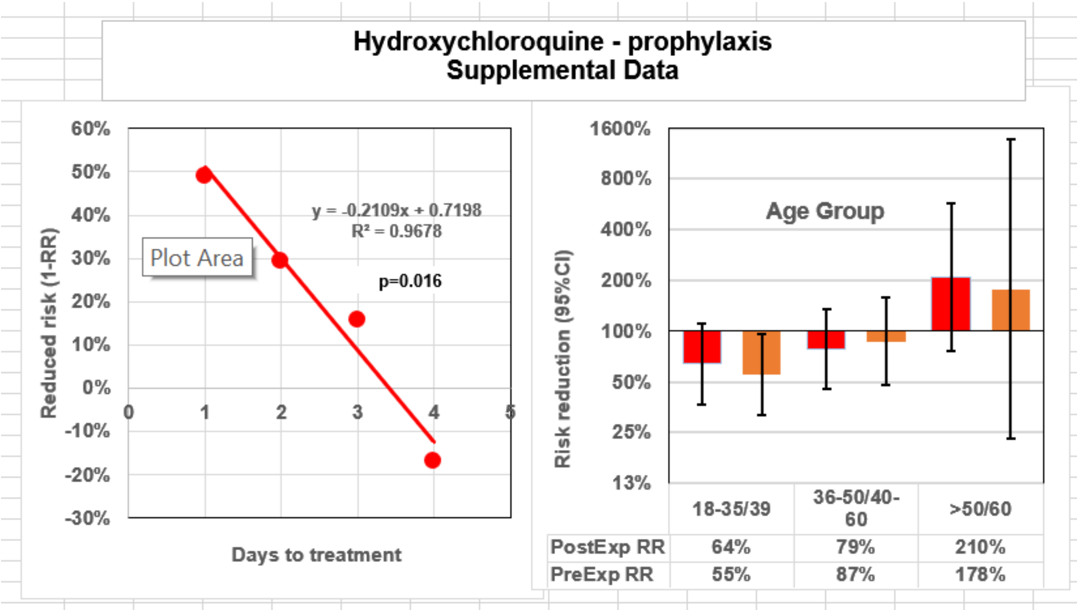


The left panel shows the effect on “treatment lag” on post-exposure prophylaxis with HCQ (based on supplemental data in Boulware[2]).The right panel shows the age stratified effect of post (red bars) or pre (brown bars) exposure prophylaxis. Note the slightly different age categories. Pre-exposure data combined from once and twice weekly HCQ treatment found in supplemental data.[1] (1 added to end point tally in >60 group to avoid division by zero error).

*Revised Understanding of the Time from Exposure to Treatment*

Our initial understanding of the Boulware paper was that the data reflected a time from exposure to treatment of up to 4 days as indicated by the concluding paragraph of the abstract:

---

[a] Our analysis, along with a number of points described in this Background section, were the subject of a letter to the New England Journal of Medicine submitted June 22 2020. Dr. Watanabe’s similar independent analysis was posted on the arxiv.org server on July 18 2020. We understand, but have not been able to confirm, that this was also submitted to the New England Journal of Medicine on June 24 2020.

*"…hydroxychloroquine did not prevent illness compatible with Covid-19 or confirmed infection when used as postexposure prophylaxis within 4 days after exposure.*

Further, the paper's discussion states:

*"…hydroxychloroquine did not prevent illness compatible with Covid-19 when initiated within 4 days after a high-risk or moderate-risk exposure."*

We were joined in our initial understanding by the distinguished company of Dr. Watanabe (who conducted an analysis similar to ours[10]), Dr. Luco (another analysis of the Boulware paper[9]), NIH (in its Treatment Guidelines,[12] p18), two Chinese organizations (in their guidelines[13]), the author of the NEJM editorial[14] accompanying the paper and the authors of a letter submitted regarding this study[15] which was not corrected by the original study authors in their reply.[5]

Not appreciated was the nuance alluded to in the paper's Table 1 ("Time from exposure to enrollment," days 1 to 4), Table S6 ("Days from Exposure"), and the extension of the eligibility window for enrollment to within 4 days after exposure, described in its methods.

In familiarizing ourselves with the publicly released study dataset after initial registration of this protocol in preparation for our re-analysis, use of the term "screening" as an apparently distinct study event from "enrollment" evoked a series of questions to the study's principal author. Based on his responses as to how time from exposure to enrollment was recorded, shipping schedules, type of FEDEX service used and shipping delays, we calculated that there is the *possibility* for considerably wide and overlapping variations in time from exposure to delivery of study drug (Table 2).

This does not take into account time zone differences, delays between enrollment and shipping, delivery delays that were experienced by some subjects and the time from receipt of study drug to time the drug was first taken. It was therefore possible that a "Day 1" subject may have initiated treatment at the same time after exposure as a "Day 4" subject. A similar issue may also have bearing on the companion study involving early treatment of COVID-19.[3]

*Table 2: Possible range of time from exposure to drug receipt, based on clarifications to released dataset*

| "Days from exposure" | Days |
|---|---|
| "Day 1" | 1.4 – 4.4 |
| "Day 2" | 2.4 – 5.4 |
| "Day 3" | 3.4 – 6.4 |
| "Day 4" | 4.4 – 7.4 |

Given that the time from exposure to treatment is arguably the central element in any study of post exposure prophylaxis, data will need to be re-stratified according to the actual individual times from exposure to delivery, which we requested. As of version 1.2 of this protocol, Dr. Boulware kindly provided the individual subject data regarding time from enrollment to receipt of study drug. Adding to this the number of days from exposure to enrollment (as per the published paper) and with certain caveats (see **Error! Reference source not found.**), these data reveal that the time from exposure to drug receipt, may extend to 6.8 days, with further uncertainty (± 24 hours) related to the unknown time that both exposure and enrollment occurred. A preliminary re-stratification reveals that HCQ when given up to 3 days post-exposure, may reduce the development of COVID-19 by as much as 65% (Table 3), in contrast to the earlier estimates of a 49% reduction within one day.

*Table 3: Effect of hydroxychloroquine on post-exposure development of COVID-19, interim estimate based on new data supplied for enrollment to delivery*

| Exposure – receipt | %pos-HCQ | %posPlac | Risk Ratio* | 95% CI |
|---|---|---|---|---|
| 0-3 days | 5.4 | 15.7 | 0.35 | 0.13 – 0.93 (p=0.044, Fisher's test) |
| 3-4 days | 12.4 | 15.3 | 0.81 | 0.4 – 1.6 |
| 4-5 days | 12.7 | 14.9 | 0.86 | 0.45 – 1.6 |
| 5-7 days | 15.7 | 11.4 | 1.37 | 0.69 – 2.7 |

Interim estimate based on additional data provided by Dr. Boulware regarding time from enrollment to drug receipt (time of first dose is unknown). Not accounting for enrollment time, the exposure-receipt time may vary $\pm$ 24 hours. These estimates do not account for time zone differences, but include a nominal 6-hour same day delivery time for Canadian subjects. n = number of subjects in each time stratum for HCQ of Placebo.

We have requested further data that will reduce the uncertainty in the estimate of exposure to drug receipt (see **Error! Reference source not found.**). Since a subject having a relatively long enrollment to delivery time likely indicates that enrollment took place earlier in the day, and vice versa for a subject with a relatively short enrollment to delivery time, this additional data may change the way subjects are stratified, attenuating the range of the RR shown in Table 3.

Further, there may be time-related biases related to the exclusion of the 100 subjects who were randomized but who became symptomatic before study drug was received. We have requested the details of these patients which are missing from the dataset, as the protocol (p26) states: *"Participants who become symptomatic with COVID19 before receiving the study medicine will be censored from the primary analysis on incident disease, but will be separately described."*

The follow-up period is similarly ambiguous, with *"outcome data being measured within 14 days after enrollment,"* when it appears that this refers to the 14th day after receipt of study drug, which could have occurred at least the next day after enrollment.

A related issue concerns the determination of the date of exposure. Dr. Boulware has provided us with the wording of the pertinent screening question as follows: *"Date of highest risk exposure. Recall that this is the day that you were closest to a COVID-19 contact, for the longest time, and with the least personal protective equipment."* Accordingly, as Dr. Boulware has pointed out in correspondence, in addition to the epidemiologically linked "index" exposure that is the target of the post-exposure prophylaxis attempted by this study, subjects may have had other exposures to multiple contacts before those contacts were diagnosed definitively or presumptively with COVID-19. Any of these other exposures alone or in aggregate may have constituted the actual "index" exposure. Given the pragmatic nature of the study, the known issues with availability and reliability of testing when the study was conducted, as well still poorly understood relationship between the amount and frequency of viral exposure and infection rate, this issue will remain a study limitation, with the estimates of time from exposure to treatment being minimum estimates. It is there likely that the subject population in this study, in terms of its actual exposure to COVID-19 this study has much in common with the population from the companion study on pre-exposure prophylaxis with HCQ[1] (see Figure 1).

*Type and Level of Exposure*

There were also differences in development of COVID-19 by type of highest exposure (HCW vs. household; (OR 0.53, CI 0.299-0.94, $p$ = 0.031) in the placebo group, with a small (8%) reduction in COVID-19 with HCQ in HCW, and a 31% reduction in the household group (RR 0.691, CI 0.398-1.2, p=0.24). The definitions of severity of exposure did not discriminate between the numbers of exposures or durations longer than 10 minutes. Differences between the different exposure types may have been accounted for by a lower age in the HCW group and higher exposure risk in household contacts, who may have had less access to advanced PPE than HCW as well as to training on hygiene practices. Differences between exposure risk related to type of in hospital-assignment and household risk among UK HCW and likely use of PPE has been suggested to account for differences in rates of COVID-19 in a UK study.[16] Despite some partial clarification, there remained some ambiguity in the revised (090920) dataset regarding the classification of risk of exposure. We have received an explanation that the description of the exposure risk score as stated in the manuscript is incorrect.

*Observational Use of Zinc and Vitamin C*

The study also included observational data relating to use of zinc and Vitamin C. There was a higher incidence of symptoms when Vitamin C was used, both without (20.8% vs 11.2%, p=0.014) and with (14.3 vs. 10.6%, p=0.33) HCQ. For zinc there was a similar relationship in the HCQ group, but not in the placebo group. Since details about timing, dose and reasons for self-medicating with these agents are unknown, these observational data confound the overall findings.

*Use of folate for placebo*

Further confounding the study is the use of folate for the placebo. Folate is considered essential for immune function.[17] In silico analyses show that folate may interact with SARS-Cov-2.[18,19] In an observational study, blood folic acid levels were significantly lower in severe COVID-19 patients.[20] For other viruses (Zika[21]; HPV[22,23]; HIV[24]) there may be an association between folate deficiency / supplementation and disease severity, amelioration or prevention.

Given the prevailing understanding of the role of endothelial dysfunction in the pathophysiology of COVID-19, restoration (or stabilization) of endothelial function may be one of several strategies to prevent or treat COVID-19. High doses (5mg, daily) of folic acid (with Vitamin B6) improves NO mediated vasodilation in children with diabetes, and has been proposed to be a useful adjunct in improving pulmonary perfusion and reducing hypoxemia in COVID-19.[25] Folate supplementation has been proposed to be a protective factor for COVID-19 in pregnant women.[26] Although the dose of folate used in the Boulware study was not described, the companion study[3] did report the strength of the folate placebo tablets in US patients as 400mcg (lactose was used for placebo in Canadian subjects). Dr. Boulware has confirmed to us that that was the case also for the post-exposure prophylaxis study. With a regime of seven tablets on the first day and 3 tablets daily thereafter, this represents folate doses of 2.8mg initially, followed by 1.2mg daily, certainly within " pharmacological range" of the doses used to improve vasodilation.[25]

Conversely, the folate receptor - FRβ - on macrophages is upregulated in inflammatory conditions[27] and may point to a negative effect of folate in COVID-19. In vitro studies found inhibition of SARS-Cov-2 replication by methotrexate,[28,29] this effect being synergistic with remdesivir and rescued by folinic acid.[28] Methotrexate, a "folate antagonist" has been proposed for use in COVID-19[30] (with folinic acid rescue[31,32]) *inter alia*, because of its effects on lymphocytes.[33] Overall changes in lymphocyte levels in COVID-19 are well described[34] with an emergingly complex picture of subset (e.g. CD8), marker expression[35] and functionality[36] changes. Folate deficiency inhibits the proliferation of CD8+ T cells in vitro.[37]

To examine the possible effect of folate, we can compare those subjects who were 100% or 0% adherent to the study drug regime from Table S6 in Boulware: (Table 4).

*Table 4: Effect of study drug adherence in post-exposure prophylaxis study*[2]

| Comparison | HCQ | Placebo | Risk Ratio | 95% CI |
|---|---|---|---|---|
| 100% adherence | 13.8% | 14.9% | 0.93 | 0.64-1.35 |
| 0% Adherence | 3.1% | 8.9% | | |

*Shown is the percent of subjects with COVID-19 positive outcome*

The aggregate COVID-19 positive outcome in subjects taking no study medication (after assignment to either group) was 5.8%, suggesting a large negative effect of the folate placebo. However, these data include:

- 11 Canadian subjects who received lactose placebo.
- 72 subjects for whom no outcome data was obtained because of Lost to Follow Up, or Withdrawal of Consent. As per the planned ITT analysis, these patients were assumed not to have achieved the study end point.

Adjusting for these subjects, the "no folate control" cohort had a slightly reduced (RR 0.93) development of COVID-19 (not statistically significant) compared with the "folate only placebo" cohort (Table 5), resulting in a small change in the estimation of the effect of HCQ in the overall study population.

*Table 5: Effect of folate placebo in post-exposure prophylaxis study*[2]

| | HCQ | | Placebo | | | |
|---|---|---|---|---|---|---|
| | **Total** | **%** | **Total** | **%** | **RR** | **95% CI** |
| HCQ vs. Folate only placebo | 47/348 | 13.5% | 51/337 | 15.1% | 0.89 | 0.82-0.97 |
| HCQ vs. no Folate control | 47/348 | 13.5% | 9/64 | 14.1% | 0.96 | 0.50-1.86 |
| No Folate control vs. Folate only Placebo | 9/64 | 14.1% | 51/337 | 15.1% | 0.93 | 0.54-1.61 |

*Shown is the percent (n/N) of subjects with COVID-19 positive outcome for the fully and partially adherent subgroups combined*

This possible negative effect of folate would also need to be considered in the other re-analyses of this study (time, contact type, use of zinc etc.) as well as in the stratifications for time and gender. The overall lower folate levels in white, African American and Mexican American men[38] as well as in Chinese men[39] may affect gender differences in folate placebo response, along with the fact that the folate dose/kg body weight in men would have been smaller.

#### 3 PRE-EXPOSURE PROPHYLAXIS (PrEP) STUDY OF HYDROXYCHLOROQUINE

This preprinted RCT ("Rajasingham," NCT04328467)[1], a companion to the Boulware study, examined the effect of once or twice weekly HCQ (800mg loading dose, then 400mg once or twice weekly for 12 weeks) on pre-exposure prophylaxis in 1483 HCW with a high exposure risk. Compared with placebo, both once (HR 0.72, 95%CI 0.44 - 1.16; p=0.18) and twice (HR 0.74, 95%CI 0.46 - 1.19; p=0.22) weekly HCQ reduced the development of COVID-19. The authors concluded that pre-exposure prophylaxis with HCQ *"once or twice weekly did not significantly reduce laboratory-confirmed Covid-19 or Covid-19-compatible illness among healthcare workers."*

As with the two companion studies,[2,3] this (likely pragmatic) study appears generally well conducted, and creatively overcame a number of logistical challenges in a short period of time. However, the powering of the study for a 50% effect size likely far exceeds lower but certainly clinically meaningful targets. This is compounded by the early termination of the study due to poor recruitment.

As with the Boulware study, the definitive nature of the study conclusion without qualification of the inadequate power, early termination and encouraging subgroup signals (mostly found in the appendix), in our opinion, impedes further investigation of promising avenues of research that may contribute significantly to mitigating the medical and economic effects of COVID-19.

Due the limitation in the Boulware study that participants were likely not subjected to a single "index" exposure, but rather to a series of exposures, there is considerable overlap in the subject populations for the pre- and post- exposure studies (but see sensitivity analyses there). Accordingly, there will likely be a number of outcome similarities. Subject to prospective confirmation, there may well be effects in specific subgroups. Although the authors combined data for the two HCQ treatment arms for the entire population (HR 0.73, CI 0.48-1.09, p=0.12), this was not done for the subgroups. However, statistically significant or near-significant effects were observed when the two arms were combined for:

- Age 18-39 (HR 0.55, CI 0.32-0.96, p=0.038) (similar to the post-exposure study, Figure 1)
    - It is unclear why in this study the age categories of 18-39, 40-60 and >60) were used, compared with those in the companion studies of 18-35, 36-50 and >50, with a much lower number of subjects in the highest age category (Table 1). For comparison,the age stratification should be rematched to the companion studies.
- Female gender (HR 0.60, CI 0.36-0.99, p=0.051)
- First responders (HR 0.46, CI 0.15-0.88, p=0.036)
- Adherent to study medication (HR 0.67, CI 0.42-1.08)

As in the companion studies, a folate placebo was used. The effect of folate, if any, is likely to be smaller than in the companion studies given only once or twice weekly dosing of smaller dose. Data for the two placebo regimes have been pooled, but should be examined separately along with the generation of a "no folate" control cohort. Although applicable to only three patients, the placebo used in Canada was stated as being a 1000mcg folate tablet, not 400mcg as in the other studies. Given the detail provided in the companion studies, this study is surprisingly silent on the possible out-of-protocol use of zinc or vitamin C.

Accordingly, a similar re-analysis of this Rajasingham study is warranted.

#### 4 TREATMENT OF EARLY COVID-19 WITH HYDROXYCHLOROQUINE

A pragmatic RCT[3] ("Skipper") which shared the same clinical trial registration (NCT04308668) as the Boulware study, examined the effect of HCQ on early COVID-19, using the same dosing (1.2g day 1 followed by 600mg daily for 4 days). Subjects not meeting the criteria for the Boulware study because they were already symptomatic were screened for the Skipper study. These patients included the 100 patients randomized in the Boulware study but who became symptomatic before receipt of study medication. There was a reduction in the percentage of subjects with ongoing symptoms at 14 days when treatment with HCQ (24%, 49/201) was compared with folate placebo (30%, 59/194) (RR 0.8, 0.58-1.1, p= 0.21). The primary end point was however a change in overall symptom severity (as a

VAS score) over 14 days. The authors concluded that HCQ *"did not substantially reduce symptom severity in outpatients with early, mild COVID-19.* This apparent inefficacy is shared by two similarly designed studies.[40,41]

Because this is a companion study to the Boulware study, it shares a number of similar issues, some of which we described in a letter submitted to the Annals of Internal Medicine and posted online.[42] These include the issue of underpowering, over-ambitious estimation of a clinically meaningful target effect size (at least for the originally intended endpoint), consideration of time, age and gender subgroups, confounding by ex-protocol and undefined use of zinc and ascorbic acid, and use of folate as placebo. To this end the reduction (Table S2) in VAS score (3.18) for the folate placebo subgroup with <75% adherence was greater than (2.15) for folate subgroup with >75% drug adherence, with intermediate values for the equivalent HCQ groups.

Patients were enrolled who had *"4 or fewer days of symptoms and either PCR-confirmed SARS-CoV-2 infection or compatible symptoms after a high-risk exposure to a person with PCR-confirmed COVID-19 within the past 14 days."* Like the companion Boulware study, the time from enrollment to delivery of study drug was likely not considered (perhaps up to 3 days) and may affect substantially the analysis and interpretation of data. Accordingly, an unknown number of patients may have initiated therapy far too late for a possible effect. The reduction in VAS score change of 26% with a duration of symptoms of 1-2 days (score difference 0.66 (95%CI -1.29 to – 0.02) was considered implausible by the authors due to the small changes for symptom onset of < 1 day (5.5%) or 3-4 days (0.2%). Although the authors suggested that differences in age and other factors may have explained this anomaly, inappropriate time stratification because enrollment to drug delivery time was not factored in, may explain this apparent anomaly.

As the initial intended endpoint of hospitalization proved inappropriate as the study progressed; a symptom-based VAS score was implemented. Unvalidated and subject to left-censoring, the use of this score obscures an apparent population bimodality ("improvers" vs. "non-improvers"). With only 24% and 30% of participants accounting for the 14-day VAS of 1.5 (HCQ) and 1.87 (placebo) respectively, average scores in still-symptomatic patients *increased* to 6.15 and 6.14 respectively.

Accordingly, we plan to subject this study to re-analysis, once the dataset becomes available.
